# Common and rare variants in HFE are not associated with Parkinson’s disease in Europeans

**DOI:** 10.1101/2020.10.22.20217190

**Authors:** Prabhjyot Saini, Sara Bandres-Ciga, Jose Luis Alcantud, Clara Ruz, Ziv Gan-Or, for the International Parkinson’s disease Genomics Consortium

## Abstract

A recent study suggested that the p.H63D variant in *HFE*, a gene involved in iron homeostasis, may modify α-synuclein pathology, the pathological hallmark of Parkinson’s disease (PD). If indeed this gene and specific variant are involved in PD, we expect to find differential distribution of *HFE* variants when comparing PD patients and controls. We analyzed genome-wide association study (GWAS) data from 14,671 PD patients and 17,667 controls and full sequencing data from additional 1,647 PD patients and 1,050 controls, using logistic regression models, and burden and Kernel tests. The *HFE* p.H63D variant was not associated with PD, nor did all the other common variants in the *HFE* locus. We did not find association of rare *HFE* variants with PD as well in all types of burden and Kernel tests. Our results do not support a role for *HFE* in PD.

## Introduction

Parkinson’s disease (PD) is a complex neurodegenerative disorder, caused in most cases by multiple genetic and environmental factors, and the normal process of aging. Previous studies have suggested that iron homeostasis may have a role in PD; pathological and imaging studies showed that iron accumulation in the substantia nigra may contribute to PD pathogenesis (Ward et al., 2014). Multiple genes are involved in iron homeostasis, one of the most important of them being *HFE*, encoding the homeostatic iron regulator protein (also called hereditary hemochromatosis protein). This protein is a membrane protein thought to regulate iron absorption, and bi-allelic *HFE* mutations may lead to hereditary haemochromatosis, an iron storage disorder.

Despite several negative genetic studies (Biasiotto et al., 2008; Duan et al., 2016; Guerreiro et al., 2006; Rhodes et al., 2014; Xia et al., 2015), a recent study suggested that the p.H63D variant in *HFE* may contribute to Parkinson’s disease pathogenesis by modifying α□synuclein pathology in cell models (Kim et al., 2020). The same group has previously suggested, using a mouse model with the homolog of the same variant (H67D), that *HFE* is a disease modifier in PD (Nixon et al., 2018). In order to examine whether this and other *HFE* variants are associated with PD, we performed a comprehensive analysis of common and rare *HFE* variants in large case-control cohorts of PD patients and controls of European origin.

## Methods

Two cohorts were included in the current study: 1) a total of 14,671 PD patients and 17,667 controls collected through collaborators from the International PD Genomics Consortium (IPDGC), and 2) 1,647 PD patients and 1,050 controls from AMP-PD (https://amp-pd.org/). Demographic details on these cohorts can be found in Supplementary Table S1A and S1B. We performed standard genome-wide association study (GWAS) quality control (QC) on the individual and variant level data as previously described (Nalls et al., 2019). We performed a similar QC process for the AMP-PD whole genome sequencing data, as detailed in the AMP-PD portal (https://amp-pd.org/whole-genome-data). Genotype data was extracted from both datasets for all variants within 100kb upstream and downstream of *HFE*, and was annotated using ANNOVAR (Wang et al., 2010). To examine whether common variants (allele frequency >0.01) in *HFE* are associated with PD, we performed logistic regression adjusted for age, sex, 10 principal components and sites of cohorts. To determine whether rare variants (allele frequency <0.01) we performed a set of burden and Kernel association tests included in the RVtests R package (Zhan et al., 2016). Bonferroni correction for multiple comparisons was applied, and all code used in the current study is available at the IPDGC GitHub at https://github.com/ipdgc/IPDGC-Trainees/blob/master/. The institutional review board (McGill University Health Center Research Ethics Board -MUHC REB) approved the study protocols.

## Results

None of the common variants in the *HFE* region was associated with risk of PD (Figure 1). The specific *HFE* variant recently reported to be associated with decreased aggregation of α□synuclein, p.H63D, had similar allele frequencies in patients and controls (0.15 and 0.14, respectively), and was not associated with risk of PD (*p*=1 after correction for multiple comparisons). Another *HFE* variant previously reported to be associated with PD, p.C282Y, had similar frequencies in patients and controls as well (0.049 in both) and was not associated with PD (corrected *p*=1). Similarly, none of the burden and Kernel association tests we used to examine the association of rare *HFE* variants with PD yielded statistically significant results (Table 1). The list of all common and rare variants found in *HFE* is available in Supplementary Table S2 and S3, respectively.

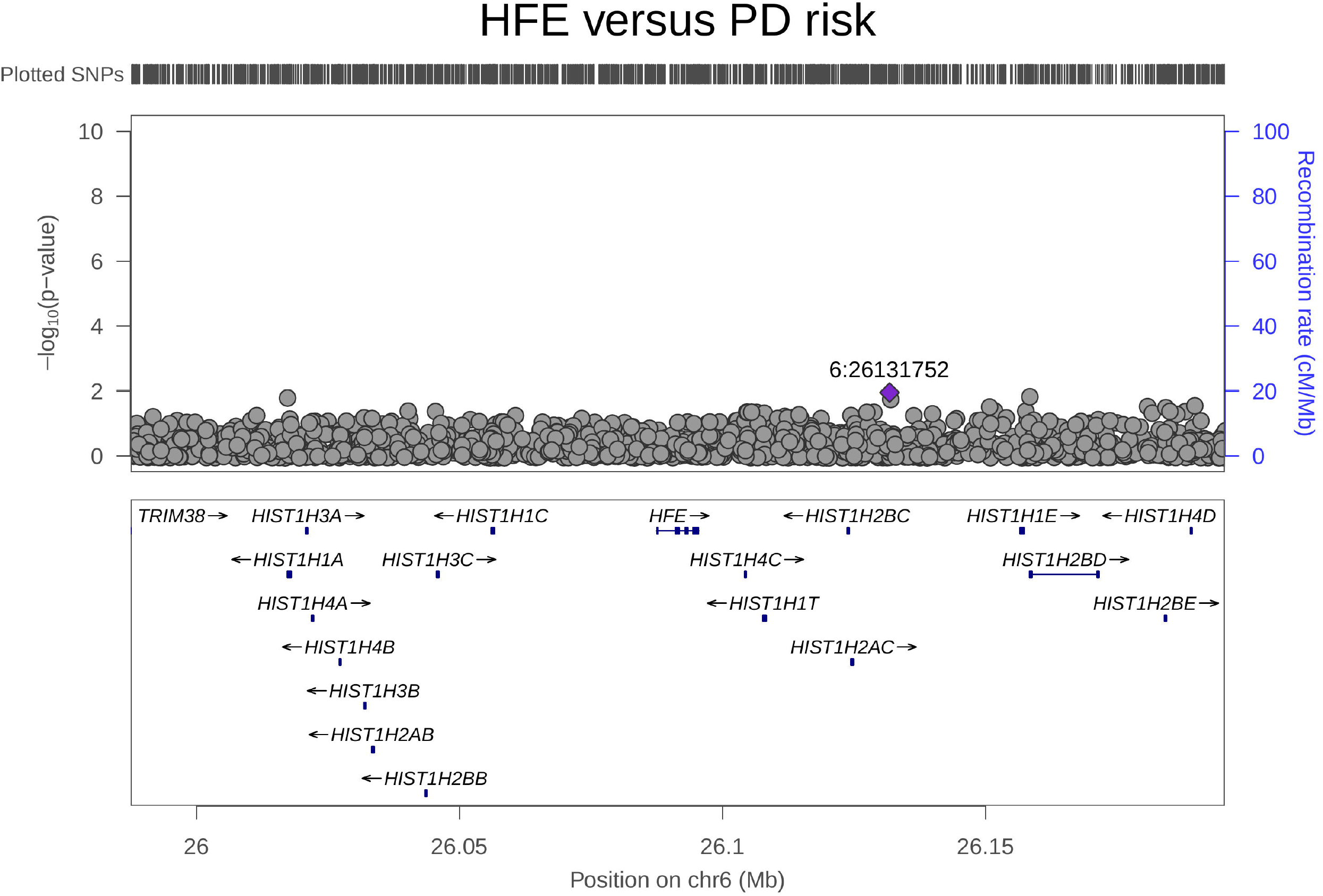

**Table 1.** Burden tests for *HFE* in the AMP-PD cohorts.

| Cohort | Burden test | All variants |  |  | Coding variants |  |  |
| --- | --- | --- | --- | --- | --- | --- | --- |
|  |  | Number of Variants | Number of Polygenic Variants | P-value | Number of Variants | Number of Polygenic Variants | P-value |
| AMP-PD | Zeggini | 54 | 47 | 0.5167 | 12 | 9 | 0.3644 |
|  | Fp | 54 | 47 | 0.4510 | 12 | 9 | 0.1134 |
|  | CMC | 54 | 47 | 0.2539 | 12 | 9 | 0.3644 |
|  | Madson-Browning | 54 | 47 | 0.2940 | 12 | 9 | 0.2959 |
|  | SKAT | 54 | 47 | 0.8299 | 12 | 9 | 0.8266 |
|  | SKAT-O | 54 | 47 | 0.6310 | 12 | 9 | 0.3425 |

Abbreviations: SKAT: Sequence kernel association test, SKAT-O: Sequence kernel association test optimized, CMC: combined multivariate and collapsing

## Discussion

In the current study we performed a thorough genetic analysis of *HFE* in large case-control cohorts of PD patients and controls. Our results do not support a role for common or rare *HFE* variants in PD. Previous smaller genetic association studies focusing on *HFE* reported contradicting results, as some studies reported associations of different *HFE* variants with PD, while other studies reported lack of association (Biasiotto et al., 2008; Borie et al., 2002; Buchanan et al., 2002; Duan et al., 2016; Guerreiro et al., 2006; Halling et al., 2008; Rhodes et al., 2014; Xia et al., 2015). Specifically for p.H63D, all the previous studies, including two meta-analyses (Duan et al., 2016; Xia et al., 2015), reported lack of association. Together with our study, there is no support from human genetic studies in PD patients for the involvement of *HFE* p.H63D in PD. Furthermore, there is no convincing evidence to suggest an association between other *HFE* variants and risk of PD. We cannot rule out that a very small effect does exist, but since it was not detected by the recent GWAS with over 50,000 PD patients and proxy-cases (Nalls et al., 2019), it is likely to be too small to be clinically meaningful, if at all.

The main limitation of the current study is that it includes only participants of European descent, and since PD is a complex disorder, it is possible that in other populations *HFE* variants may still contribute to the disease when found on another genetic background. In a recent GWAS of Asian population, however, no GWAS-significant association has been reported for *HFE* (Foo et al., 2020).

Despite the lack of association in the current study, the role of iron and iron homeostasis in PD should be further studied, whether by genetic studies or by biochemical, functional, clinical and pathological studies. We recommend that genetic data from the IPDGC and other available studies should be consulted before studying specific genetic variants in different models.

## Data Availability

Data used for the analysis can be found in the supplementary tables. The code used for the analysis is available at https://github.com/ipdgc/IPDGC-Trainees

## Acknowledgements

We thank the participants for contributing to the study. We would like to also thank all members of the International Parkinson Disease Genomics Consortium (IPDGC). For a complete overview of members, acknowledgements and funding, please see http://pdgenetics.org/partners. This research was supported in part by the Intramural Research Program of the NIH, National institute on Aging. This work was also supported in part by grants from the Michael J. Fox Foundation, the Canadian Consortium on Neurodegeneration in Aging (CCNA), and the Canada First Research Excellence Fund (CFREF), awarded to McGill University for the Healthy Brains for Healthy Lives initiative (HBHL). ZGO is supported by the Fonds de recherche du Québec - Santé (FRQS) Chercheurs-boursiers award, in collaboration with Parkinson Quebec, and by the Young Investigator Award by Parkinson Canada.

